# NMR metabolomic profiles in white British and British Indian vegetarians and non-vegetarians in the UK Biobank

**DOI:** 10.1101/2025.10.03.25337243

**Authors:** Qiaoye J. Wang, Julie A. Schmidt, Timothy J. Key, Fiona Bragg, Diego Aguilar-Ramirez, Vivian Viallon, Ruth C. Travis, Tammy Y.N. Tong

## Abstract

**Background:** The human metabolome is influenced by diet, and studying circulating metabolites may help clarify underlying mechanisms linking diet groups and disease outcomes, but few large studies have been conducted. This study investigated differences in plasma metabolites across diet groups in the UK Biobank.

**Methods:** The UK Biobank recruited 500,000 adults (aged 40-69) across the UK in 2006-2010. At recruitment, participants reported ethnicity and diet, from which we defined six diet groups in white British participants (regular meat eaters, low meat eaters, poultry eaters, fish eaters, vegetarians, vegans) and two in British Indian participants (meat eaters, vegetarians). Metabolomics profiling (249 plasma metabolites) was performed using nuclear magnetic resonance (NMR) in a random subset of ∼275,000 participants. We used multivariable-adjusted linear regression to estimate differences in adjusted geometric means of metabolite levels by diet group.

**Results:** Significant differences between diet groups were observed in 241 (97%) metabolites in white British participants after multiple testing correction. Compared with regular meat eaters, vegetarians and vegans had lower concentrations of n-3, DHA, and their ratios to total fatty acids. However, they had higher n-6, linoleic acid, and their ratios to n-3 and total fatty acids. Additionally, non-meat eaters had higher glycine but lower branched-chain amino acid concentrations. Large differences were also seen in many lipoprotein subclasses. Similar patterns were observed in British Indian participants.

**Conclusions:** Marked differences in metabolite profiles across diet groups signify variations in fatty acid, amino acid, and lipid intake and metabolism, which may help explain associations with long-term health outcomes.

**Highlights:**

- People on different diets showed substantial differences in metabolomic profiles.
- Largest differences were seen in several fatty acids and their ratios.
- Similar patterns were found in white British and British Indian populations.

## 1 Introduction

Previous studies have shown differences in many disease outcomes in people with varying degrees of animal food exclusion. For example, vegetarians and vegans have been observed to have lower risk of ischaemic heart disease [1], diabetes [2], and possibly some cancers compared with omnivores [3], but higher risk of haemorrhagic stroke [1] and fractures [4,5]. These differences in disease risk may be preceded by changes in relevant biomarkers that reflect disease metabolism or mechanisms. Indeed, previous studies have identified differences in several established biomarkers. For example, lower concentrations of low-density lipoprotein cholesterol (LDL-C) and insulin-like growth factor 1 (IGF-1) have been observed in both vegetarians and vegans in a previous UK Biobank observational study, with the lowest concentrations seen in vegans [6]. These biomarker differences contribute to understanding the observed differences in cardiovascular and cancer risk, respectively [7,8]. However, the underlying mechanisms linking vegetarian diets with most disease outcomes are not well established, and further studies on diet and disease-related biomarkers are warranted.

In contrast to the assessment of individual biomarkers, metabolomics enables the simultaneous measurement of a wide range of circulating metabolites in the human body, which could help enhance our understanding of metabolic processes and disease mechanisms [9]. However, limited evidence is available on how metabolomic profiles may vary by diet group, and existing studies are comparatively small in sample size, with the largest published studies including fewer than 100 vegetarians or vegans [10]. Therefore, further evidence from large-scale studies is needed to clarify any differences in metabolite concentrations by diet group. Additionally, dietary choices can differ substantially between ethnic groups, and genetic predisposition may influence metabolism and nutrient processing [11]; yet it remains unknown whether any metabolomic differences observed by diet group would be consistent across different ethnic groups, as previous studies have focused primarily on populations of white European ancestry.

The aim of this study is to provide a detailed description of circulating metabolite concentrations and ratios by diet group in white British and British Indian participants with varying degrees of animal food exclusion, using data from the UK Biobank, a large population-based cohort in the United Kingdom with metabolomics data available for approximately 275,000 individuals.

## 2 Methods

### 2.1 Study Population

The UK Biobank is a prospective cohort study that recruited 500,000 middle-aged people (aged 40-69 years) from across the United Kingdom (including England, Wales and Scotland) in 2006 to 2010. The scientific rationale and design of the UK Biobank have been described in detail previously [12]. Briefly, participants were identified from National Health Service registers, and were invited to participate in the study if they resided within travelling distance (∼25 km) of one of the 22 assessment centres. At baseline, the participants attended an assessment centre where they completed a touchscreen questionnaire asking about lifestyle (including diet, alcohol consumption, smoking status, physical activity), socio-demographic characteristics, and general health and medical history. All participants also participated in a verbal interview, and had their physical measurements and blood samples taken by trained staff. The UK Biobank study was approved by the National Information Governance Board for Health and Social Care and the National Health Service Northwest Multicentre Research Ethics Committee (06/MRE08/65), and all participants gave informed consent to participate using a signature capture device at the baseline visit [12].

### 2.2 Ethnicity classification

On the touchscreen questionnaire, participants were asked to self-identify their ethnicity as ‘White’, ‘Mixed’, ‘Asian or Asian British’, ‘Black or Black British’, ‘Chinese’, ‘Other ethnic group’, ‘Do not know’, or ‘Prefer not to answer’, with further sub-categories under each option. For the current analyses, participants were included if they self-identified as ‘White’, or as ‘Asian or Asian British’ and subsequently as ‘Indian’. These groups are hereafter referred to as ‘white British’ and ‘British Indian’. The white British population was included as it constituted the majority of the UK Biobank population (94%), while the British Indian population was included because of its relatively large proportion of vegetarians (26%). Participants of other ethnic groups were excluded from the analyses due to the small numbers of non-meat eaters, which limited valid comparisons by diet group.

### 2.3 Diet group classification

Participants were classified into diet groups based on self-reported dietary information from the touchscreen questionnaire completed at recruitment. On the questionnaire, participants were asked about their frequency of consumption of processed meat, beef, lamb or mutton, pork, poultry (such as chicken or turkey), oily fish, and other types of fish, with the possible responses ranging from “never” to “once or more daily”. Participants were also asked whether they never consumed dairy and eggs or foods containing eggs. Based on their responses to these questions, the white British participants were classified into six diet groups: regular meat eaters (red and processed meat consumption >3 times per week), low meat eaters (red and processed meat consumption ≤3 times per week), poultry eaters (participants who ate poultry but no red and processed meat), fish eaters (participants who ate fish, but not red and processed meat and poultry), vegetarians (participants who did not eat any meat or fish, but ate one or both of dairy or eggs), and vegans (participants who did not eat any meat, fish, dairy or eggs). The British Indian participants were classified into two diet groups, meat eaters (ate any combination of red or processed meat or poultry) and vegetarians; because the numbers of fish eaters and vegans among the British Indians were small (each fewer than 100) and their metabolomic profiles may be distinct, these two diet groups were neither defined separately nor combined with other diet groups in the analyses of this population.

### 2.4 Metabolites measurement

Non-fasting blood samples were collected from all UK Biobank participants by trained personnel (either a phlebotomist or a nurse), except in a small proportion (0.3%) of participants who declined, were deemed unable to donate a sample, or where the attempt was unsuccessful due to technical or health reasons. Metabolomics assays were performed in a random subsample of 275,000 participants from baseline recruitment in two phases: ∼118,000 participants in phase 1 and ∼157,000 participants in phase 2. Details of the sample preparation, assay procedures, and other technical details have been described elsewhere [13–15]. In brief, circulating metabolites were measured in ethylenediaminetetraacetic acid (EDTA) plasma samples, using Nightingale Health’s nuclear magnetic resonance (NMR)-based metabolic biomarker profiling platform (Nightingale Health Ltd, Helsinki, Finland). Samples were prepared directly in 96-well plates by UK Biobank and shipped on dry ice to Nightingale Health’s laboratories for analyses.

The Nightingale Health platform uses a targeted high-throughput NMR spectrometry approach to quantify 249 metabolite measures (168 measures in absolute levels and 81 ratio measures) in a single assay. This includes biomarkers related to lipid and cholesterol metabolism (including 14 lipoprotein subclasses), fatty acid compositions, and various low-molecular weight molecules such as amino acids, ketones and glycolysis metabolites, as well as markers of fluid balance and inflammation. Each lipoprotein subclass was characterised in terms of its triglycerides, phospholipids, total cholesterol, cholesterol esters, free cholesterol, and total lipid concentrations.

For clarity in results presentation, metabolites were classified into eight groups based on their metabolic class or functions: amino acids; cholesterol, free cholesterol & cholesterol esters; fatty acids and ratios; glycolysis, ketone bodies, fluid balance, inflammation; lipids, phospholipids, triglycerides & apolipoproteins; lipoprotein particle properties; lipoprotein subclasses; and relative lipoprotein lipid concentrations.

### 2.5 Inclusion and exclusion criteria

Participants were excluded from the analyses if they had no data on any metabolites (n=227,809, 274,132 remained). We further excluded participants who were not of white or British Indian ethnicity (n=11,760), did not answer a sufficient number of questions to be classified into one of the pre-specified diet groups (n=1,944), or had missing data on fasting time (n=5). Given that most of the measured metabolites were lipid metabolism related, we also excluded all participants who reported taking lipid-lowering medications at baseline, based on responses from the verbal interview (n=44,419). A participant selection flowchart is shown in **Supplementary Figure 1**.

### 2.6 Statistical analyses

Baseline characteristics of UK Biobank participants included in the analyses were tabulated by six diet groups among the white British participants, and by two diet groups among the British Indians. For statistical analyses, all metabolite variables were first logarithmically transformed to prepare for estimation of geometric means. Multivariable-adjusted linear regression models were then fitted with the log-transformed metabolite concentrations or ratios as the outcome and diet group as the main exposure, adjusting for the covariates sex, age (5-year categories), BMI (in kg/m^2^; <20, 20.0-22.4, 22.5-24.9, 25.0-27.4, 27.5-29.9, 30.0-32.4, 32.5-34.9, ≥35, unknown), alcohol consumption (<1, 1-7, 8-15, ≥16 g/day, unknown), smoking status (never, previous, current <15 cigarettes/day, current ≥15 cigarettes/day, unknown), physical activity (low <10, moderate 10-49.9, high ≥50 excess metabolic equivalent of task hours/week, unknown), geographical region (London, North-West England, North-East England, Yorkshire, West Midlands, East Midlands, South-East England, South-West England, Wales, Scotland), fasting time (0-1, 2, 3, 4, 5, 6-7, ≥8 hours), and spectrometer number (1-8) for the NMR assays.

Adjusted geometric mean concentrations and ratios of metabolites across diet groups were estimated using predictive margins from multivariable-adjusted linear regression models. These margins represent the predicted mean for each diet group, standardised to the same covariate distribution. The predicted values on the log scale were exponentiated to obtain the final adjusted geometric means. Percent differences in adjusted geometric means of metabolite concentrations and ratios between each diet group and regular meat eaters in white British (or meat eaters in British Indian) were calculated. Wald tests were used to assess heterogeneity (overall and pairwise comparisons with regular meat eaters) across the six diet groups in the white British population and the two diet groups in the British Indian population. Post-estimation linear combinations of parameters were used to compare vegetarians and vegans in the white British population. To account for multiple testing while allowing for correlations between the exposures, we conducted a principal component analysis of the metabolite variables, and determined that the first 49 (white British) and 43 (British Indian) principal components explained 99% of the total variation in the exposure data [16]. Consequently, the effective numbers of independent tests were determined to be 49 and 43. The statistical significance level after Bonferroni correction for multiple testing was thus set to 0.05/49=0.0010 and 0.05/43=0.0012 for white British and British Indian, respectively.

In addition to being a potential confounder, BMI may also act as a mediator between diet and metabolite concentrations. Therefore, to examine the influence of BMI on the observed associations, we conducted a sensitivity analysis excluding BMI from the covariates. All analyses were performed using Stata version 18.5 (StataCorp, TX, USA). Figures were generated using “ggplot2” package version 3.5.1 and “Jasper” package version 2.266 in R version 4.4.2 [17].

## 3 Results

### 3.1 Baseline characteristics

The analyses included a total of 216,004 participants, comprising 213,944 white British and 2,060 British Indian participants. **Table 1** presents the baseline characteristics of the study population by ethnicity and diet group. Among white British participants, vegetarians and vegans were generally younger and more likely to be women compared to regular meat eaters. They were also less likely to be overweight or obese, consumed less alcohol, were less likely to be current smokers, were more physically active, and were less likely to have a history of diabetes. Among British Indian participants, vegetarians were older, more likely to be women, had marginally higher prevalence of overweight or obesity, reported less alcohol consumption, had a lower proportion of current smokers, and had a higher prevalence of diabetes compared to meat eaters. No significant difference in physical activity was observed between British Indian diet groups. In both ethnic groups, a slightly higher proportion of non-meat eaters were assessed in London than in the other regions. Fasting duration did not differ meaningfully across diet groups in either ethnic group.

**Table 1.**
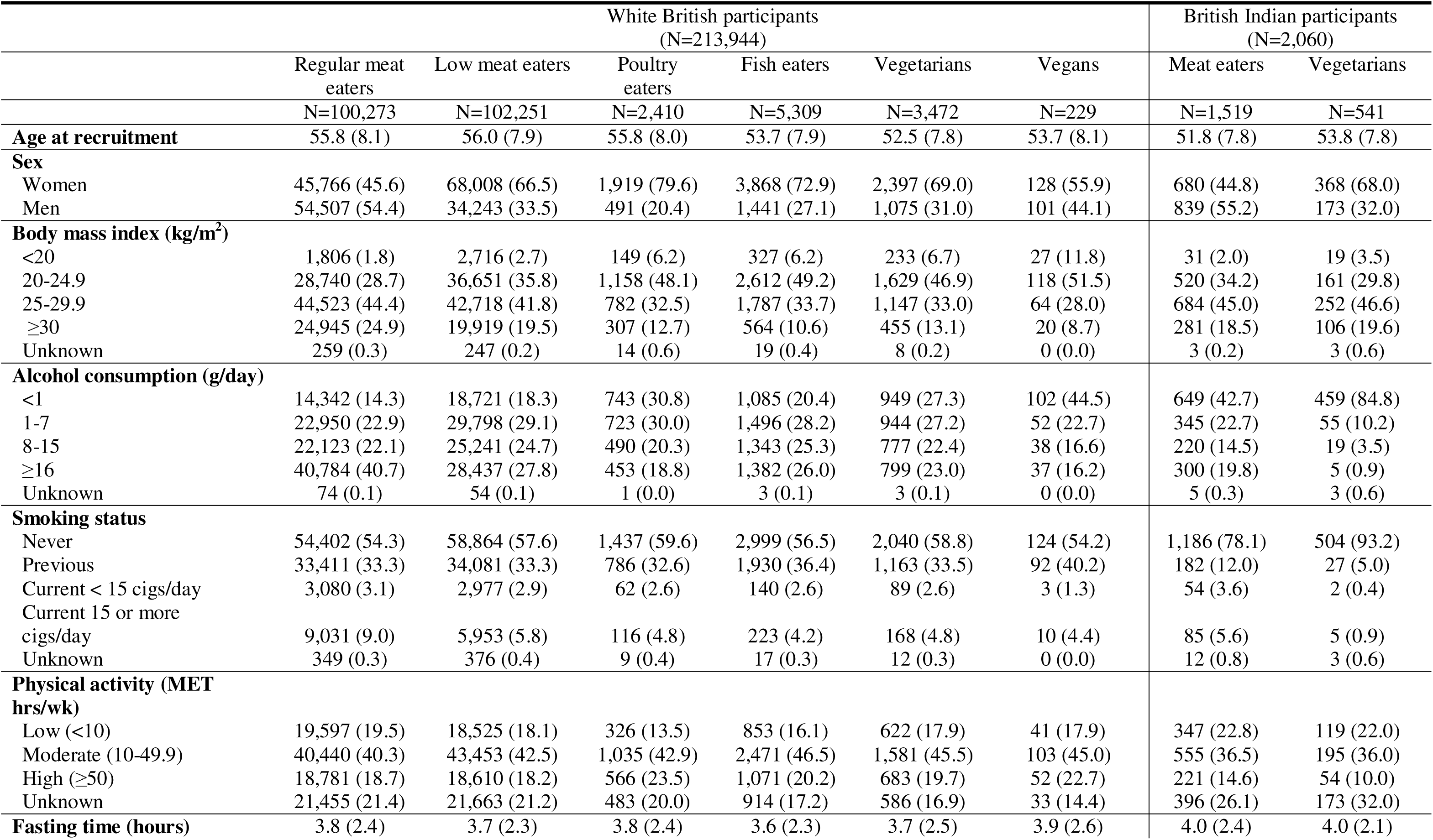
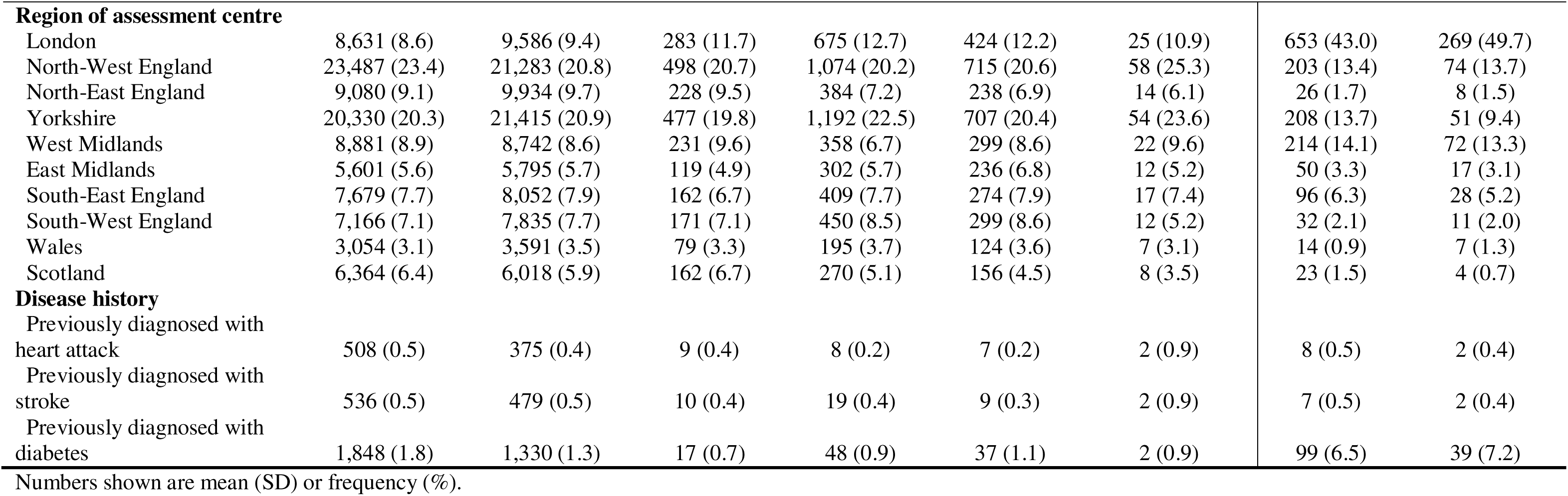
Baseline characteristics of white British and British Indian participants by diet groups in UK Biobank (N=216,004)

### 3.2 Metabolic profiles by diet group

Significant differences by diet group were observed for 241 (97%) metabolites in white British participants and 23 (9%) in British Indian participants after correction for multiple testing. Although the number of statistically significant findings differed, likely due to differences in statistical power between the two populations, the directions and overall patterns of metabolite differences by diet group were broadly consistent in the two ethnic groups. Pairwise comparisons between each diet group and regular meat eaters (white British participants) and between vegetarians and meat eaters (British Indian participants) are presented as volcano plots. **Figure 1** shows vegetarians versus the reference group in both ethnic groups. Remaining diet groups versus regular meat eaters, plus vegans versus vegetarians in white British participants, are shown in **Supplementary Figures 2-6**. Complete results of all significantly different metabolites and their corresponding percent differences are provided in **Supplementary Table 1**.

**Figure 1.**
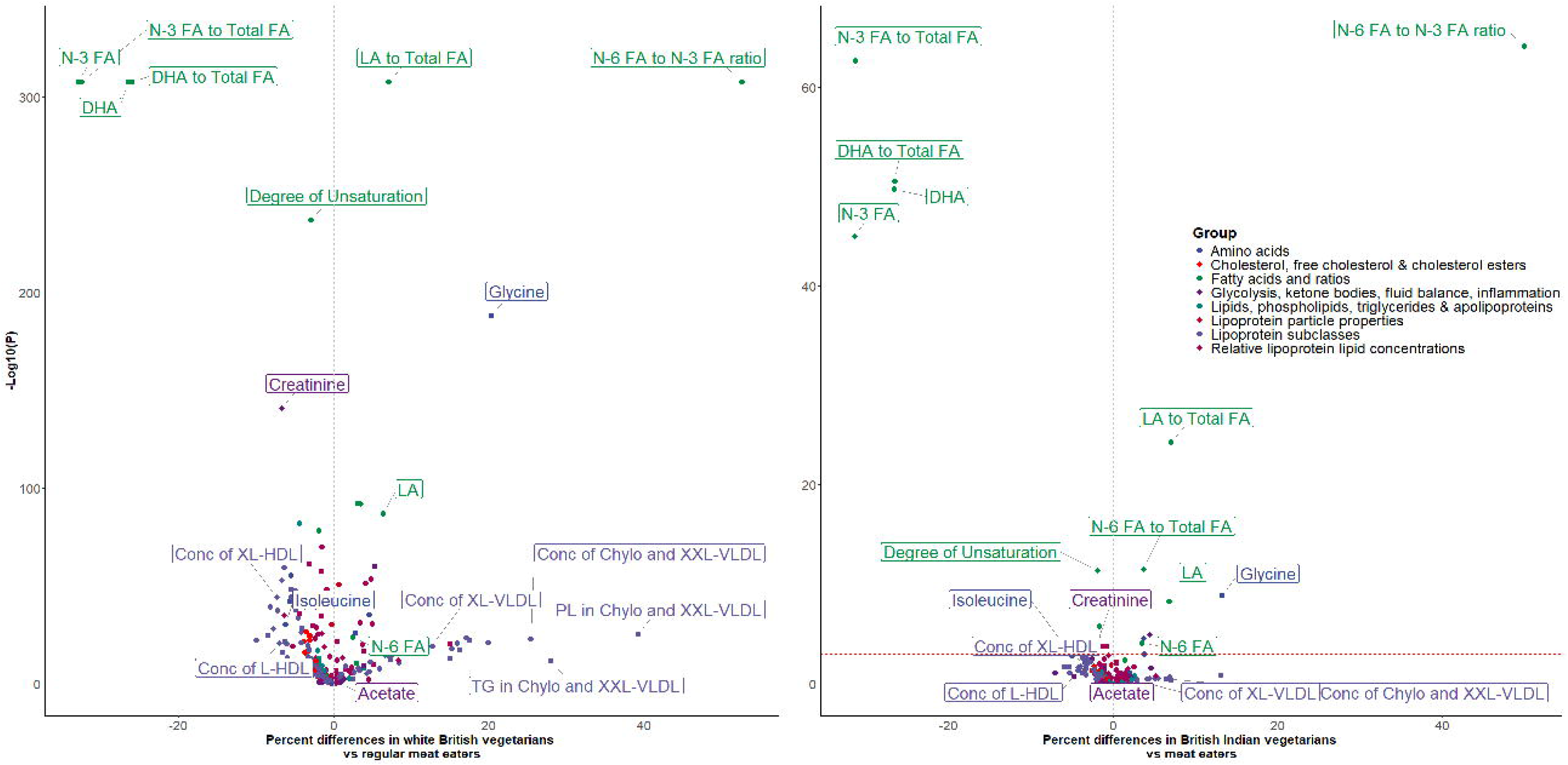
Volcano plots of metabolites comparing vegetarians versus regular meat eaters in white British and vegetarians versus meat eaters in British Indian participants Caption: The red dashed line indicates the statistical significance threshold after Bonferroni correction for multiple testing based on the effective number of independent tests (*p* < 0.0010 for white British and *p* < 0.0012 for British Indian participants). The grey dashed vertical line marks zero percent difference between diet groups. Metabolites were labelled if they met any of the following criteria: 1). Extremely small p-values (< 1X10^−100^ for white British and < 1X10^−10^ for British Indian participants); 2). Large absolute percent difference (> 20%) and statistically significant; 3). Selected for detailed comparisons across diet groups in subsequent figures. Abbreviations: Chylo: chylomicrons; Conc: concentration; DHA: docosahexaenoic acid; HDL: high-density lipoproteins; L: large; LA: linoleic acid; MUFA: monounsaturated fatty acids; N-3 FA: n-3 fatty acids; N-6 FA: n-6 fatty acids; PL: phospholipids; TG: triglycerides; VLDL: very low-density lipoproteins; XL: very large; XXL: Extremely large.

Comparing vegetarians with regular meat eaters or meat eaters, metabolites in the fatty acids and their ratios group showed the largest percent differences and smallest p-values (**Figure 1**). Many lipid subclass metabolites, including concentrations of chylomicrons & extremely large-VLDL, as well as a number of other metabolites such as glycine and isoleucine, also showed large differences between vegetarians and meat eaters.

To further illustrate the differences observed, a subset of metabolites with the most pronounced differences between non-meat eaters and meat eaters was selected and plotted across all diet groups in both ethnicities in **Figures 2-5**.

**Figure 2.**
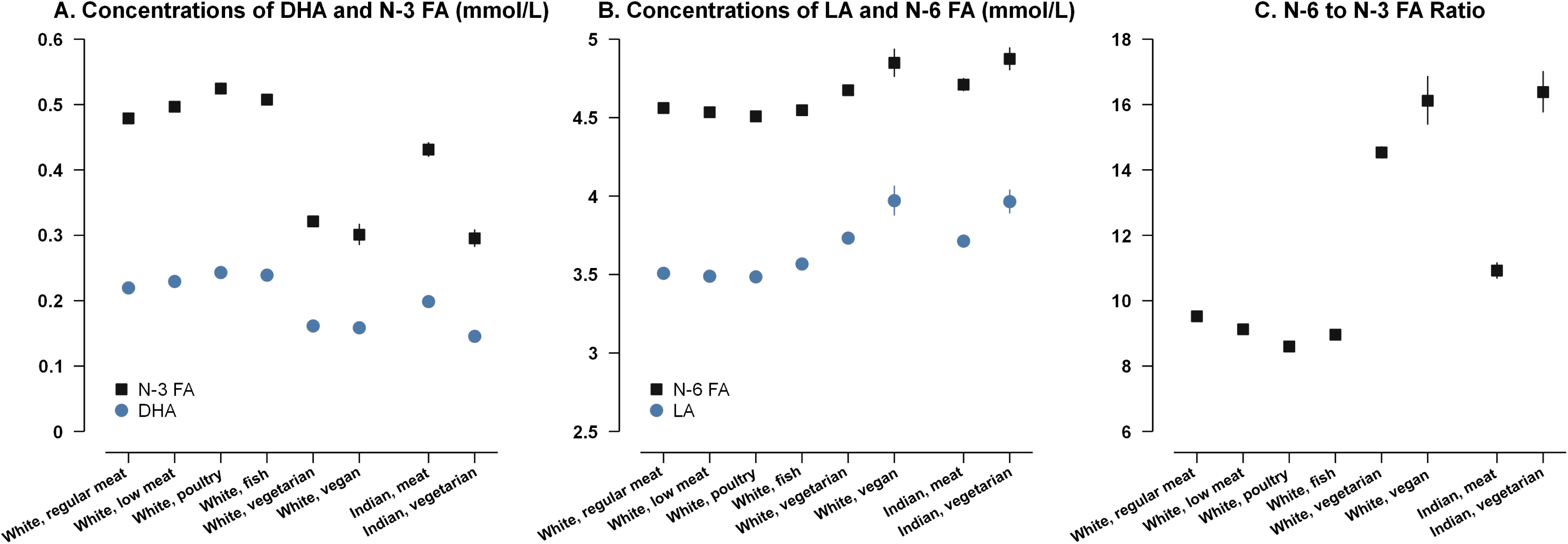
Selected fatty acid and ratio concentrations by diet group and ethnicity in the UK Biobank Caption: Point estimates represent adjusted geometric mean levels (95% CIs) estimated from multivariable linear regression models, adjusted for sex, age, BMI, alcohol consumption, smoking status, physical activity, geographical region, fasting status, and spectrometer number. *P* for heterogeneity across the diet groups were all statistically significant after Bonferroni correction for multiple testing based on the effective number of independent tests for each ethnic groups (*p* < 0.0010 for white British and *p* < 0.0012 for British Indian participants). DHA: docosahexaenoic acid; LA: linoleic acid; N-3 FA: n-3 fatty acid; N-6 FA: n-6 fatty acid.

Compared with regular meat eaters, low meat, poultry, and fish eaters had higher concentrations of n-3 fatty acid (n-3 FA) and docosahexaenoic acid (DHA), whereas vegetarians and vegans had lower levels of these fatty acids. Specifically, in white British participants, the adjusted geometric mean concentration (95% CI) of total n-3 FA was 0.321 (0.317, 0.326) mmol/L in vegetarians and 0.301 (0.286, 0.317) mmol/L in vegans, compared to 0.479 (0.478, 0.480) mmol/L in regular meat eaters. However, vegetarians and vegans had higher concentrations of n-6 fatty acid (n-6 FA), linoleic acid (LA), and higher n-6 to n-3 FA ratios compared to regular meat eaters. Similar patterns were observed in British Indian participants.

Regarding lipoprotein subclass metabolites, vegetarians had higher concentrations of chylomicrons & extremely large-VLDL and very large-VLDL, as well as increased levels of lipid components carried within them, compared to regular meat eaters or meat eaters (**Figure 3; Supplementary Table 1**). However, for vegans in the white British group, these lipoprotein subclass metabolites showed a mixed directional pattern compared with regular meat eaters, with some showing positive and others negative percent differences. None of these differences, however, was statistically significant. On the other hand, concentrations of very large-HDL, large-HDL, and their associated lipids were found to be lower in both vegetarians and vegans compared with regular meat eaters in white British, and lower in vegetarians compared with meat eaters in British Indian participants.

**Figure 3.**
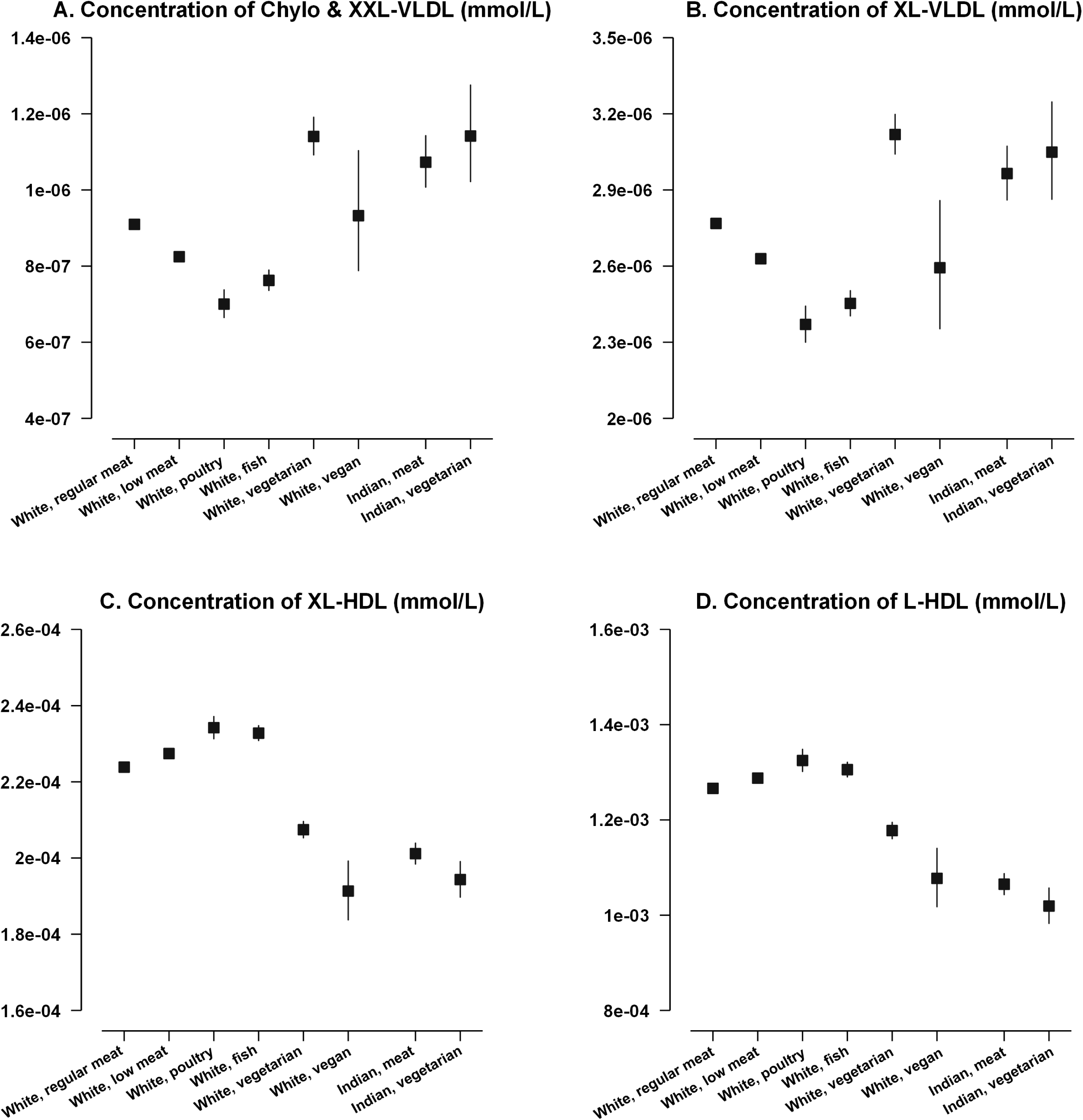
Selected lipoprotein subclass concentrations by diet group and ethnicity in the UK Biobank Caption: Point estimates represent adjusted geometric mean levels (95% CIs) estimated from multivariable linear regression models, adjusted for sex, age, BMI, alcohol consumption, smoking status, physical activity, geographical region, fasting status, and spectrometer number. *P* for heterogeneity across diet groups were all statistically significant in white British participants but not in British Indian participants, after Bonferroni correction for multiple testing based on the effective number of independent tests (*p* < 0.0010 and *p* < 0.0012, respectively). Chylo: Chylomicron; L: Large; XL: Very large; XXL: Extremely large.

For amino acids, gradient trends were observed across diet groups with increasing exclusion of animal food products from the diet in both ethnic groups. In particular, in the white British population, concentrations of glutamine, alanine, and glycine showed a gradient from lowest in regular meat eaters to highest in vegans (**Supplementary Table 2**). Glycine showed the largest percent difference between vegetarians [0.193 (0.190, 0.195) mmol/L] and regular meat eaters [0.160 (0.160, 0.160) mmol/L]: a 20.3% difference. In contrast, concentrations of total and individual branched-chain amino acids (BCAAs) [i.e., isoleucine, leucine, and valine] showed the opposite gradient, with highest concentrations in regular meat eaters and lowest in vegans. Among the BCAAs, isoleucine showed the greatest difference across diet groups, with concentrations of 0.0482 (0.0481, 0.0483) mmol/L in regular meat eaters, 0.0452 (0.0447, 0.0457) mmol/L in vegetarians, and 0.0435 (0.0417, 0.0453) mmol/L in vegans (**Figure 4)**. For the other amino acids measured, including tyrosine, histidine, and phenylalanine, no statistically significant differences were found between vegetarians and regular meat eaters.

**Figure 4.**
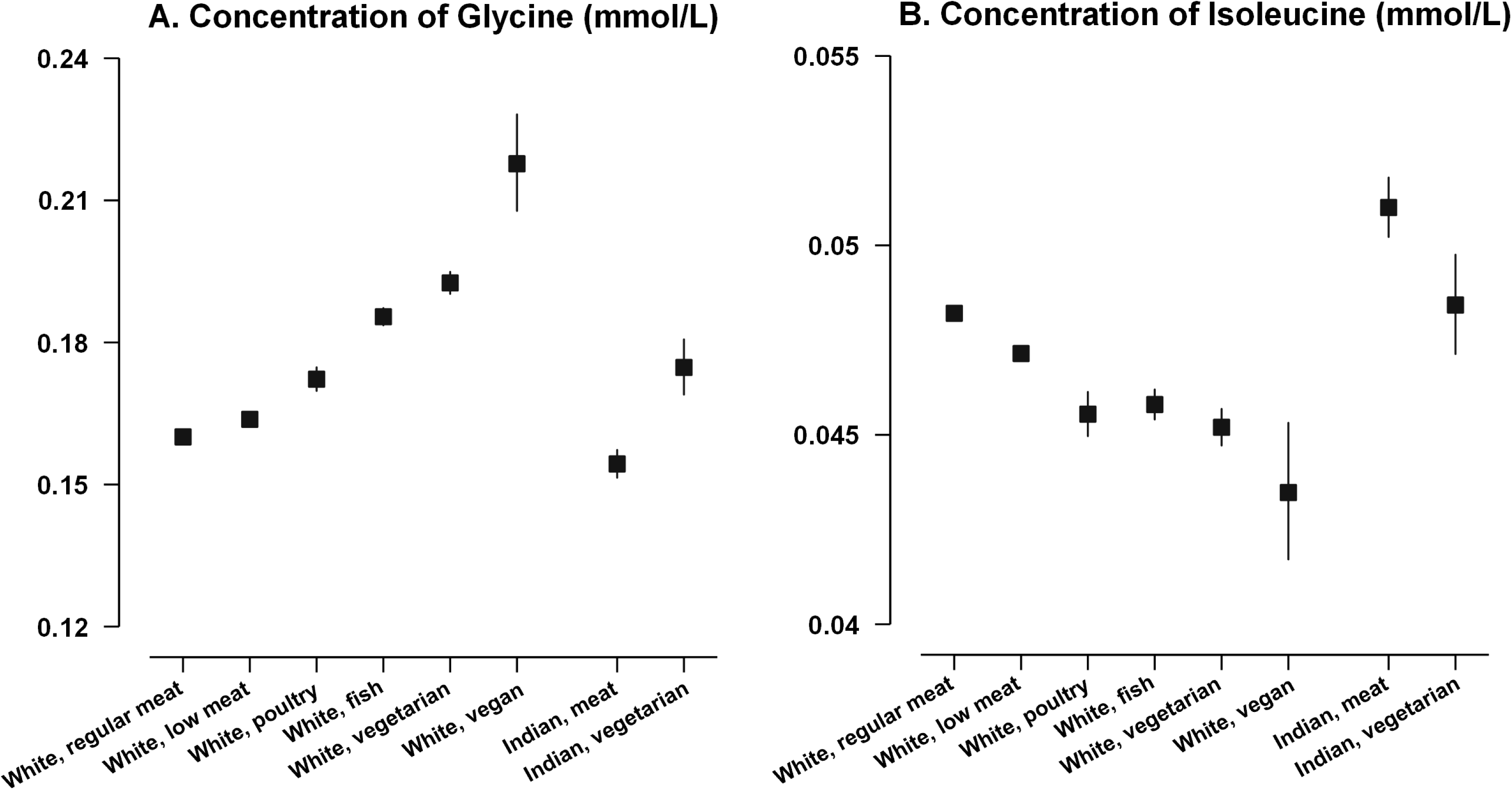
Concentrations of glycine and isoleucine by diet group and ethnicity in the UK Biobank Caption: Point estimates represent adjusted geometric mean levels (95% CIs) estimated from multivariable linear regression models, adjusted for sex, age, BMI, alcohol consumption, smoking status, physical activity, geographical region, fasting status, and spectrometer number *P* for heterogeneity across diet groups were statistically significant for both metabolites in white British participants, but only for glycine in British Indian participants, after Bonferroni correction for multiple testing based on the effective number of independent tests (*p* < 0.0010 and *p* < 0.0012, respectively).

In the group of fluid balance and ketone bodies, creatinine concentrations were gradually lower with greater exclusion of animal products (**Figure 5**). For acetate, a substantial difference in its concentration was observed between vegetarians and vegans. Vegetarians showed no difference in acetate concentration [0.0148 (0.0146, 0.0151) mmol/L] compared to regular meat eaters [0.0147 (0.0147, 0.0148) mmol/L], whereas vegans had a substantially higher estimated concentration of 0.0169 (0.0158, 0.0180) mmol/L than both vegetarians and all other diet groups (**Figure 5; Supplementary Figure 6**).

**Figure 5.**
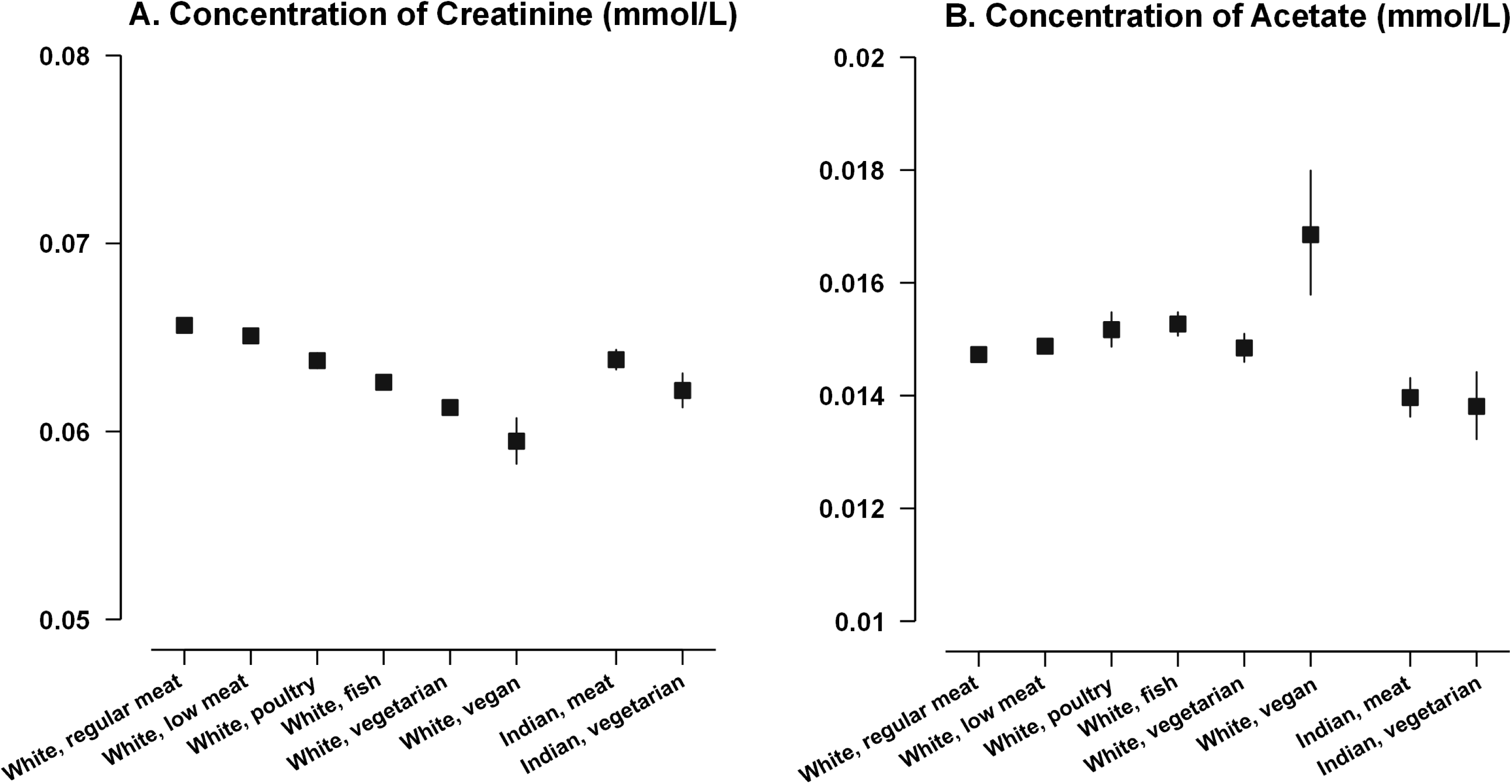
Concentrations of creatinine and acetate by diet group and ethnicity in the UK Biobank Caption: Point estimates represent adjusted geometric mean levels (95% CIs) estimated from multivariable linear regression models, adjusted for sex, age, BMI, alcohol consumption, smoking status, physical activity, geographical region, fasting status, and spectrometer number. *P* for heterogeneity across diet groups were statistically significant for both metabolites in white British participants, but not in British Indian participants, after Bonferroni correction for multiple testing based on the effective number of independent tests (*p* < 0.0010 and *p* < 0.0012, respectively).

Despite similar patterns of difference by diet group in white British and British Indian participants, substantial differences in the absolute concentrations of many metabolites were seen between participants of the same diet group in the two ethnic groups. For example, compared with white British vegetarians, British Indian vegetarians had higher concentrations of n-6 FA, LA (**Figure 2B**), and isoleucine (**Figure 4B**), but lower n-3 FA, DHA (**Figure 2A**), very large-HDL and large-HDL concentrations (**Figure 3C & 3D**), and similar concentrations of chylomicrons & extremely large-VLDL and very large-VLDL (**Figure 3A & 3B**).

In sensitivity analysis not adjusting for BMI in the model, findings for most metabolites remained largely unchanged. However, differences in lipoprotein subclass metabolites between non-meat eaters and omnivores were generally smaller, and in some cases, reversed direction. For instance, the percent difference in very large-VLDL concentrations between vegetarians and regular meat eaters in white British participants changed from 12.7% (in the model including BMI) to −2.8% (in the model without BMI) (**Supplementary Tables 2 & 3**).

## 4 Discussion

In this British cohort of ∼275,000 participants, 249 circulating metabolites and their ratios were quantified using an NMR metabolomics platform and compared across diet group and ethnicity. We observed significant differences in the concentrations of 241 (97%) metabolites in white British and 23 (9%) in British Indians by diet group, with mixed directionality depending on specific metabolites. Although fewer metabolites showed statistically significant differences in British Indians (likely due to a smaller sample size), the directions of difference comparing vegetarians with meat eaters were similar between the two ethnic groups for all metabolites. Many of the largest differences were found in metabolites in the fatty acids and their ratios group, followed by those in the lipoprotein subclasses group. Clear differences were also shown in other metabolites including a few amino acids and metabolites related to glycolysis, ketone bodies, fluid balance, and inflammation.

### 4.1 Comparison with previous studies

Few studies have examined metabolomic profiles by diet group, and previous studies have been relatively small and used a variety of metabolomics platforms [10,18–20]. For example, a study by Lindqvist et al. examined ^1^H-NMR profiling in 120 healthy adults (45 men and 75 women), including 40 omnivores/meat eaters, 13 vegetarians adding fish, 24 vegetarians, and 43 vegans [19]. The study measured ∼70 metabolites and found significantly higher concentrations of isoleucine, valine, creatine, and creatinine, and lower level of lysine, in meat eaters compared to non-meat eaters. These findings, except for lysine, which was not measured in the current assay, were also observed in our current study. Another previous study conducted in the EPIC-Oxford cohort investigated metabolic profile differences in 379 men in the diet groups of meat eaters, fish eaters, vegetarians, and vegans, using a targeted mass-spectrometry metabolomic assay (Biocrates) [18]; 93 of 118 metabolites were found to differ significantly by diet group. Due to differences in metabolomics platforms, only metabolites in the amino acids group were comparable to the current study, and this previous study also showed that glycine concentration was highest in vegans, whereas BCAAs including leucine and valine were lower in vegans.

The most comparable study to the current one was another previous EPIC-Oxford cohort study, which examined NMR metabolomic profiles of 207 metabolites by four diet groups in a small sample of men (80 meat eaters, 69 fish eaters, 74 vegetarians, 63 vegans) [10]. That study reported significant differences in fatty acid concentrations and ratios, including total n-3 FA, DHA, total n-6 FA, LA, and their corresponding ratios, as well as in a number of amino acids, creatinine, and all lipid-related traits in lipoprotein subclasses, by diet group. Findings of the current study were largely in accordance with this previous evidence.

Additionally, to our knowledge, no previous studies have examined metabolomic profile differences by diet group across two ethnic groups. The current study addressed this gap and found similar patterns of metabolite differences by diet group in both white British and British Indian participants. However, absolute concentrations of many metabolites differed between the two ethnic groups within similar diet groups. These absolute differences may be related to distinct eating behaviours and culturally influenced practices, such as cooking methods and traditional recipes, as well as genetic differences that potentially influence metabolic pathways and nutrient processing in the two populations [11,21].

Comparing to the previous studies, the current study is the largest to date investigating differences in metabolomic profiles across diet groups in two ethnic groups. We identified substantial differences in several specific metabolites, particularly between non-meat eaters and omnivores. The following sections elaborate on these specific differences.

### 4.2 Fatty acids and ratios

Vegetarians and vegans showed substantial differences in several circulating fatty acids and related ratios, including significantly higher concentrations of n-6 FA and its primary type LA, and their respective ratios to total FA, but lower concentrations of n-3 FA, DHA, and their ratios to total FA. These differences are likely attributable to dietary differences between meat and non-meat eaters, as moderately high correlations have been found between FA intakes and their circulating levels, particularly for long-chain polyunsaturated FA [22].

LA is predominantly found in plant-sourced foods such as vegetable oils and nuts. As vegetarians and vegans typically consume greater amounts of these foods than omnivores [23], it is expected that they show higher circulating concentrations of LA and n-6 FA. Conversely, the main dietary sources of the long-chain n-3 FA, eicosapentaenoic acid (EPA) and DHA, are seafood, particularly oily fish, which are totally excluded from vegetarian and vegan diets. It has been estimated that amounts of EPA and DHA consumed from dietary sources in vegans are negligible, and vegetarians consume minimal EPA (<5 mg/day) and varying but small amounts of DHA depending on egg consumption (averaging <33 mg/day) [24]. In contrast, omnivores have an average intake of 100-150 mg/day of EPA and DHA combined. For vegans, with essentially zero dietary intake, the circulating EPA and DHA levels observed reflect endogenous synthesis from alpha-linolenic acid (ALA), an essential n-3 FA mainly found in plant-based foods such as flaxseeds, walnuts, rapeseed oil, and soybeans. However, it is known that the conversion efficiency in humans is low [25]. Moreover, this conversion may be further limited by a high LA intake in vegetarians and vegans, since the desaturation of ALA and LA involves the same rate-limiting desaturase enzyme [26]. These FA findings are consistent with previous smaller studies examining FA profiles across diet groups [27,28].

### 4.3 Lipoprotein subclasses and their lipid compartments

Our results showed that white British vegetarians, but not vegans, had significantly higher concentrations of certain lipoprotein particles, particularly chylomicrons & extremely large-VLDL and very large-VLDL, compared with regular meat eaters. Conversely, concentrations of very large-HDL and large-HDL were found to be lower in both white British vegetarians and vegans than in regular meat eaters.

The observed variations in lipoprotein subclasses across diet groups were consistent with findings from the previous smaller EPIC-Oxford study, although specific estimates and statistical significance differed [10]. Dietary differences between diet groups can at least partly explain these differences. For example, the inclusion and varying intakes of dairy and eggs in vegetarians could introduce different types and compositions of fats compared to both omnivores and vegans, potentially influencing lipid-related metabolism and leading to distinct lipoprotein profiles. A previous UK Biobank study has reported that vegetarians had relatively higher dietary intakes of cheese, yoghurt, and eggs than regular meat eaters, whereas vegans had negligible consumption of these foods [29].

Nonetheless, the higher concentrations of chylomicrons & extremely large and large VLDL observed in vegetarians appeared to be influenced by BMI adjustment. Sensitivity analysis showed that these positive associations were substantially smaller, or even reversed to negative, when BMI was excluded from the model. Because non-meat eaters typically have lower BMI, the attenuations observed in the model not adjusting for BMI suggest that BMI differences may mask underlying variations in these lipoprotein subclasses between diet groups. Chylomicrons and VLDL are known as triglycerides (TG)-rich lipoprotein particles. Although the percent difference in total TG between white British vegetarians and regular meat eaters was smaller than that shown for chylomicrons & extremely large VLDL and large VLDL, it followed a similar pattern: concentration was highest in vegetarians, but the associations reversed after BMI was excluded from the covariates. A comparable pattern of lower TG before BMI adjustment but higher after adjusting for BMI in vegetarians versus meat eaters was also reported in one previous UK Biobank study examining clinical chemistry biomarkers [6].

### 4.4 Amino acids

#### 4.4.1 Glycine

Glycine is a non-essential amino acid that can be synthesised endogenously as well as obtained exogenously from dietary sources. Major dietary sources include animal products such as meat and fish [30], although some plant-based foods, such as soya products and other legumes, are also relatively rich in glycine [23]. Differences in endogenous synthesis or metabolism across diet groups may also contribute to the varying concentrations observed in this study. Our finding of higher circulating glycine concentrations in vegetarians and vegans is consistent with two previous observational studies: one conducted within the EPIC-Oxford cohort and another in a small cohort of 36 vegans and 36 omnivores in Germany [31,32]. Interestingly, these studies reported a combination of lower dietary glycine intake but higher plasma glycine levels in vegetarians and vegans, which suggested potential metabolic differences between meat eaters and non-meat eaters.

#### 4.4.2 Isoleucine

Isoleucine, together with leucine and valine, comprises the group of branched-chain amino acids (BCAAs). BCAAs are essential amino acids, with primary dietary sources including meat, fish, dairy products, and eggs [33]; plant-based sources include soybeans, chickpeas, seeds, and tree nuts [34]. However, most plant-source proteins contain relatively lower amounts of BCAAs compared with animal-source proteins [35,36]. This relatively lower BCAA content in plant foods likely contributes to the lower circulating concentrations of these amino acids observed in vegetarians and vegans in our study. This finding is consistent with previous small-scale studies (with fewer than 100 participants in each diet group), which also reported lower circulating BCAA levels, particularly in vegans, compared with omnivores [31,37].

### 4.5 Glycolysis, ketone bodies, fluid balance, inflammation

#### 4.5.1 Creatinine

Vegetarians and vegans had significantly lower creatinine concentrations than regular meat eaters, with concentration lowest in vegans. Serum creatinine is a by-product of creatine metabolism, and creatine is primarily found in protein-rich animal foods such as red meat and fish [38], which are not consumed by vegetarians and vegans. Our finding is consistent with a previous UK Biobank cohort study that measured both serum and urinary creatinine concentrations using clinical biomarker assays and found both to be lower in vegetarians and vegans compared with regular meat eaters [6].

#### 4.5.2 Acetate

Acetate is one of the most abundant short-chain fatty acids (SCFAs), which are end products of microbial fermentation of indigestible carbohydrates, such as dietary fibre, by gut bacteria [39]. Previous clinical trials have shown that high dietary fibre intake is associated with increased concentrations of plasma SCFAs, including acetate [40,41]. Vegetarians and vegans typically consume more dietary fibre [42], which therefore may lead to increased acetate production. However, in the current study, vegetarians did not show significantly different acetate concentrations compared with omnivores, whereas vegans had a significantly higher concentration than both omnivores and vegetarians. Existing studies comparing metabolomic profiles by diet group, however, did not have acetate in their measured metabolites [10,18–20]. Therefore, the particular elevated concentration of acetate observed in vegans in this study is a novel finding that, to our knowledge, has not been reported previously.

One possible explanation is that vegan diets, being more restrictive than vegetarian diets, may include higher intakes of fruits, vegetables, and legumes, all of which are rich in fermentable fibre and could promote greater acetate production [23]. Additionally, circulating acetate can be derived from endogenous metabolism through processes related to energy derivation, storage, and utilisation, involving the activities of acetyl-CoA hydrolase and synthetase [43]. It is possible that vegetarians and vegans differ in aspects of endogenous acetate metabolism due to physiological adaptations to long-term dietary patterns, although this requires further investigation.

### 4.6 Strengths and limitations

The key strength of this study is that it is the largest study to date to present metabolomic profile differences with quantification of a broad range of metabolites across diet groups of varying degrees of animal food exclusion. The inclusion of both white British and British Indian populations allowed for detailed comparisons by both diet group and ethnicity. However, several limitations should be acknowledged. First, the sample size for British Indian participants was relatively small, which limited the ability to apply the same diet group categorisations as in the white British population and reduced the statistical power for analyses in this group. Nevertheless, to our knowledge, this remains the largest multi-ethnic UK population cohort with metabolomics data. Second, some self-selection bias might be present in this cohort, as only a modest proportion (5.5%) of invited participants agreed to take part [12]. This may also limit the generalisability of the current findings to the general UK population. Nonetheless, the validity of exposure-outcome associations is not necessarily dependent on the representativeness of the participants. Third, differences in metabolite concentrations may arise from variations in sample preparation and processing, including differences in participants’ fasting times and in the spectrometers used for NMR assays [14]. However, both fasting time and spectrometer number were adjusted for in the current analyses, and any such variations are expected to be non-differential by diet group; therefore, the potential influence on the observed associations is likely minimal. Fourth, although the analyses have adjusted for several key confounders, residual confounding by other non-dietary factors may still be present. Lastly, due to the cross-sectional nature of the study, we were unable to establish causal relationships between diet group and metabolite levels, although for certain metabolites such as DHA, alternative non-dietary explanations are less likely.

## 5 Conclusions

In this large population-based UK cohort, many differences in circulating metabolites were observed between diet groups, particularly between vegetarians or vegans and omnivores, in both white British and British Indian populations. These variations may reflect not only dietary differences but also potential physiological differences associated with varying degrees of animal food exclusion, which may help explain previously reported associations between diet group and health outcomes. Further research is warranted to investigate underlying biological mechanisms linking diet group, metabolic profiles, and disease risks.

## Data Availability

All data produced in the present study are available upon reasonable request to the UK Biobank data.

## Abbreviations

ALA: Alpha-linolenic acid
BCAAs: Branched-chain amino acids
DHA: Docosahexaenoic acid
EPA: Eicosapentaenoic acid
HDL: High-density-lipoprotein
LA: Linoleic acid
NMR: Nuclear magnetic resonance
N-3 FA: N-3 fatty acids
N-6 FA: N-6 fatty acids
TG: Triglycerides
VLDL: Very-low-density-lipoprotein

## Author Contributions

The study was conceived and designed by TYNT, JAS, TJK, and RCT. TYNT and QJW analysed the data and wrote the first draft of the manuscript. All other authors provided input on data analysis and interpretation of results, and revised the manuscript critically for important intellectual content. All authors read and approved the final manuscript.

## Conflict of Interest

The authors declare no conflict of interest.

## Funding

This work was supported by UK Research and Innovation (MR/X032809/1) and Cancer Research UK (C8221/A29017 and C8221/A29186). The funders had no role in study design, data collection, analysis, decision to publish, or preparation of the manuscript. For the purpose of open access, the authors have applied a Creative Commons Attribution (CC BY) license to any Author Accepted Manuscript version arising.

## Disclaimer

Where authors are identified as personnel of the International Agency for Research on Cancer / World Health Organization, the authors alone are responsible for the view expressed in this article and they do not necessarily represent the decisions, policy, or views of the International Agency for Research on Cancer / World Health Organization.

## Acknowledgements

This research used the UK Biobank resource (application number 67506). We thank the participants of the UK Biobank for their contribution to the resource.

## Notes

### Competing Interest Statement

The authors have declared no competing interest.

### Author Declarations

The National Information Governance Board for Health and Social Care and the National Health Service Northwest Multicentre Research Ethics Committee gave ethical approval for this work.

