## Supplementary Figure 1 for "NMR metabolomic profiles in white British and British Indian vegetarians and non-vegetarians in the UK Biobank"

**Supplementary Figure 1**. Participant selection flow chart of the study

501,941 UKB participants

Excluded 227,809 participants had no data on any metabolites

274,132 participants

Excluded 11,760 participants of other (i.e. not white British or British Indian) or unknown ethnicities

Excluded 1,944 participants who could not be classified into one of the prespecified diet groups

Excluded 5 participant with missing information on fasting time

Excluded 44,419 participant reported taking lipid lowering drugs

216,004 participants, including
213,944 white British and
2,060 British Indian

**Supplementary Figure 2.** Volcano plots of metabolites in white British low meat eaters compared with regular meat eaters

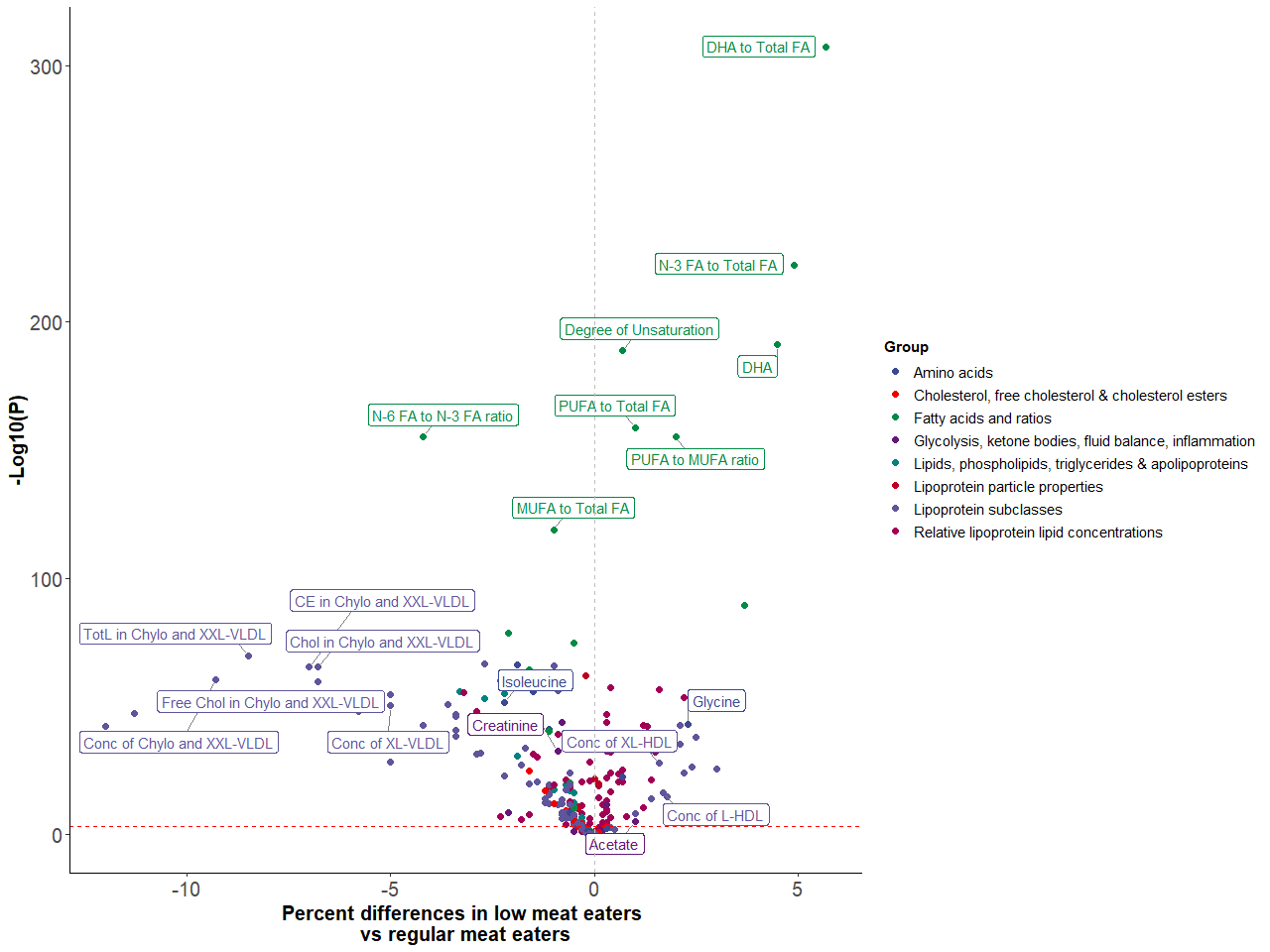

The red dashed line indicates the statistical significance threshold after Bonferroni correction for multiple testing based on the effective number of independent tests (*p* < 0.0010). The grey dashed vertical line marks zero percent difference between diet groups. Chylo: chylomicrons; Conc: concentration; DHA: docosahexaenoic acid; HDL: high-density lipoproteins; L: large; LA: linoleic acid; MUFA: monounsaturated fatty acids; N-3 FA: omega-3 fatty acids; N-6 FA: omega-6 fatty acids; PL: phospholipids; TG: triglycerides; VLDL: very low-density lipoproteins; XL: very large; XXL: Extremely large.

**Supplementary Figure 3.** Volcano plots of metabolites in white British poultry eaters compared with regular meat eaters

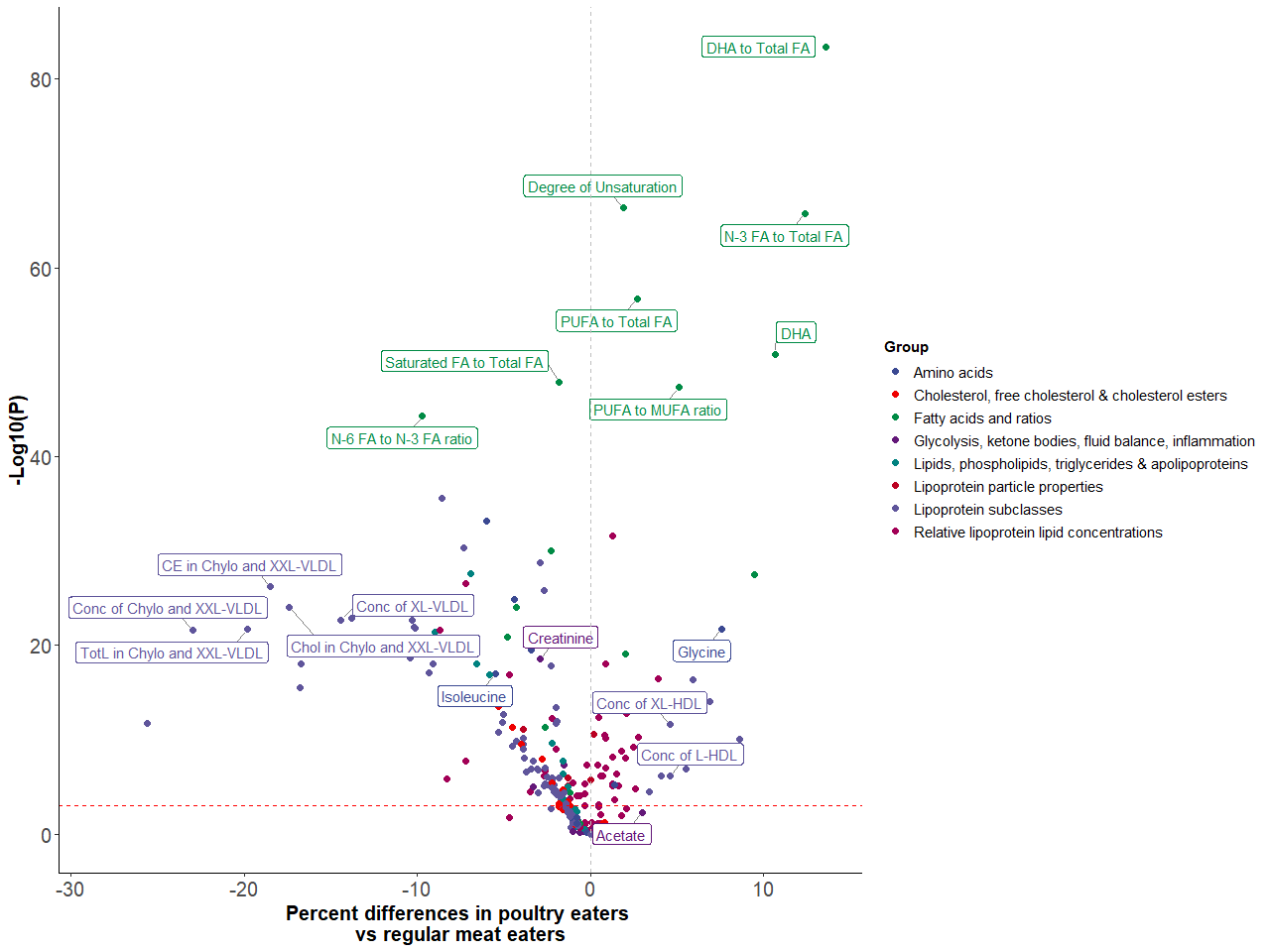

The red dashed line indicates the statistical significance threshold after Bonferroni correction for multiple testing based on the effective number of independent tests (*p* < 0.0010). The grey dashed vertical line marks zero percent difference between diet groups. Chylo: chylomicrons; Conc: concentration; DHA: docosahexaenoic acid; HDL: high-density lipoproteins; L: large; LA: linoleic acid; MUFA: monounsaturated fatty acids; N-3 FA: omega-3 fatty acids; N-6 FA: omega-6 fatty acids; PL: phospholipids; TG: triglycerides; VLDL: very low-density lipoproteins; XL: very large; XXL: Extremely large.

**Supplementary Figure 4.** Volcano plots of metabolites in white British fish eaters compared with regular meat eaters

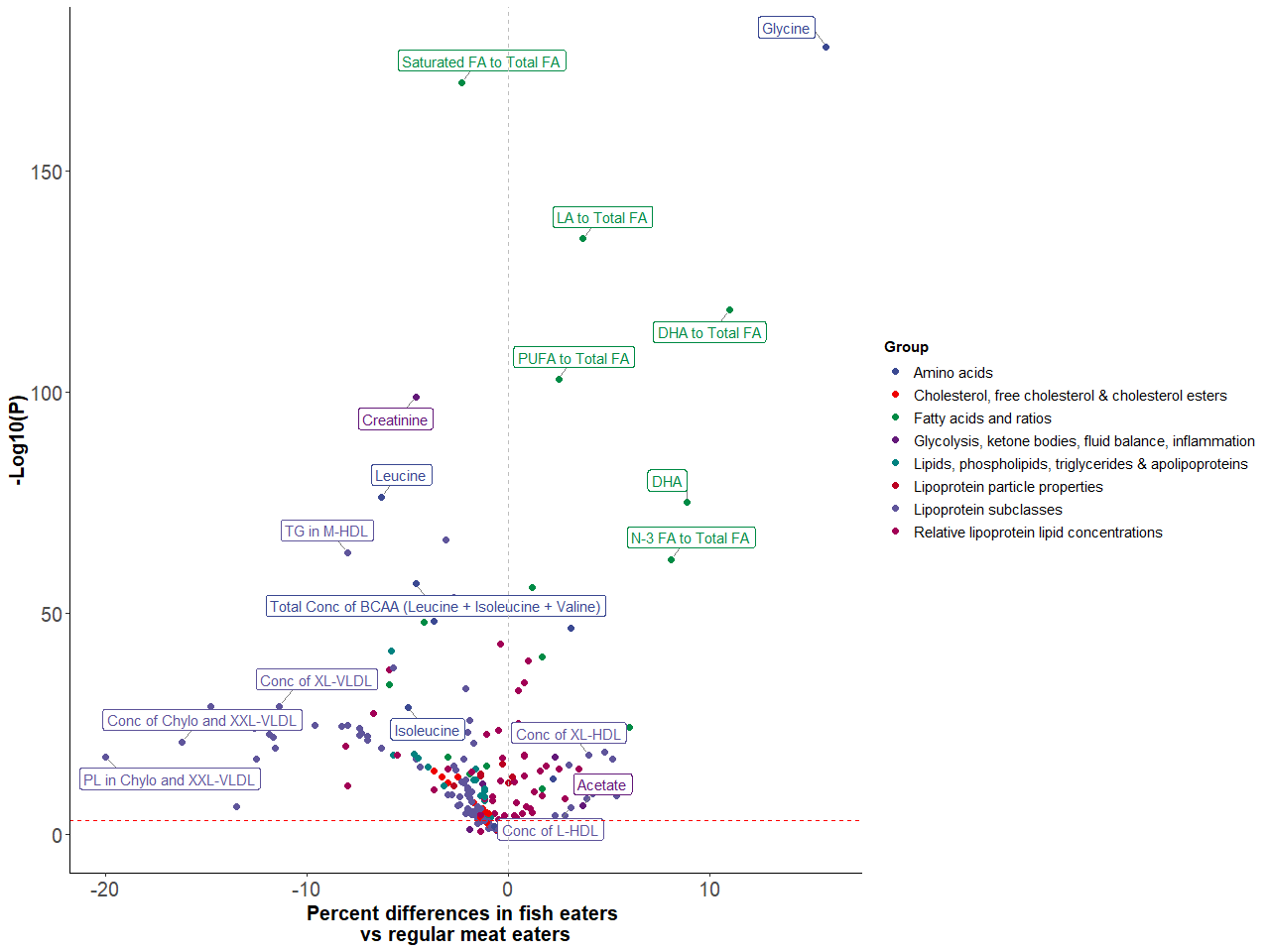

The red dashed line indicates the statistical significance threshold after Bonferroni correction for multiple testing based on the effective number of independent tests (*p* < 0.0010). The grey dashed vertical line marks zero percent difference between diet groups. Chylo: chylomicrons; Conc: concentration; DHA: docosahexaenoic acid; HDL: high-density lipoproteins; L: large; LA: linoleic acid; MUFA: monounsaturated fatty acids; N-3 FA: omega-3 fatty acids; N-6 FA: omega-6 fatty acids; PL: phospholipids; TG: triglycerides; VLDL: very low-density lipoproteins; XL: very large; XXL: Extremely large.

**Supplementary Figure 5.** Volcano plots of metabolites in white British vegans compared with regular meat eaters

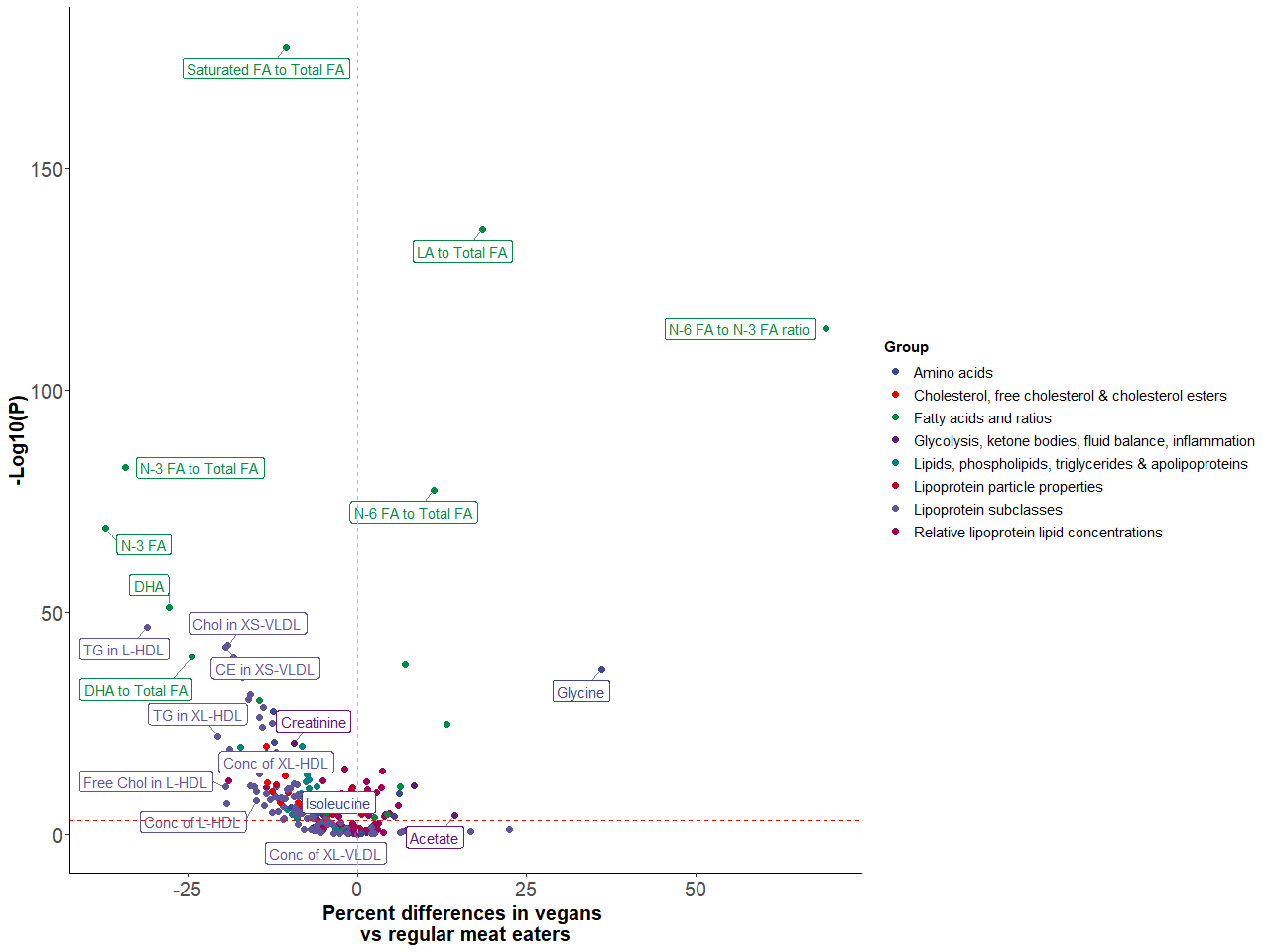

The red dashed line indicates the statistical significance threshold after Bonferroni correction for multiple testing based on the effective number of independent tests (*p* < 0.0010). The grey dashed vertical line marks zero percent difference between diet groups. Chylo: chylomicrons; Conc: concentration; DHA: docosahexaenoic acid; HDL: high-density lipoproteins; L: large; LA: linoleic acid; MUFA: monounsaturated fatty acids; N-3 FA: omega-3 fatty acids; N-6 FA: omega-6 fatty acids; PL: phospholipids; TG: triglycerides; VLDL: very low-density lipoproteins; XL: very large; XXL: Extremely large.

**Supplementary Figure 6.** Volcano plots of metabolites in white British vegans compared with vegetarians

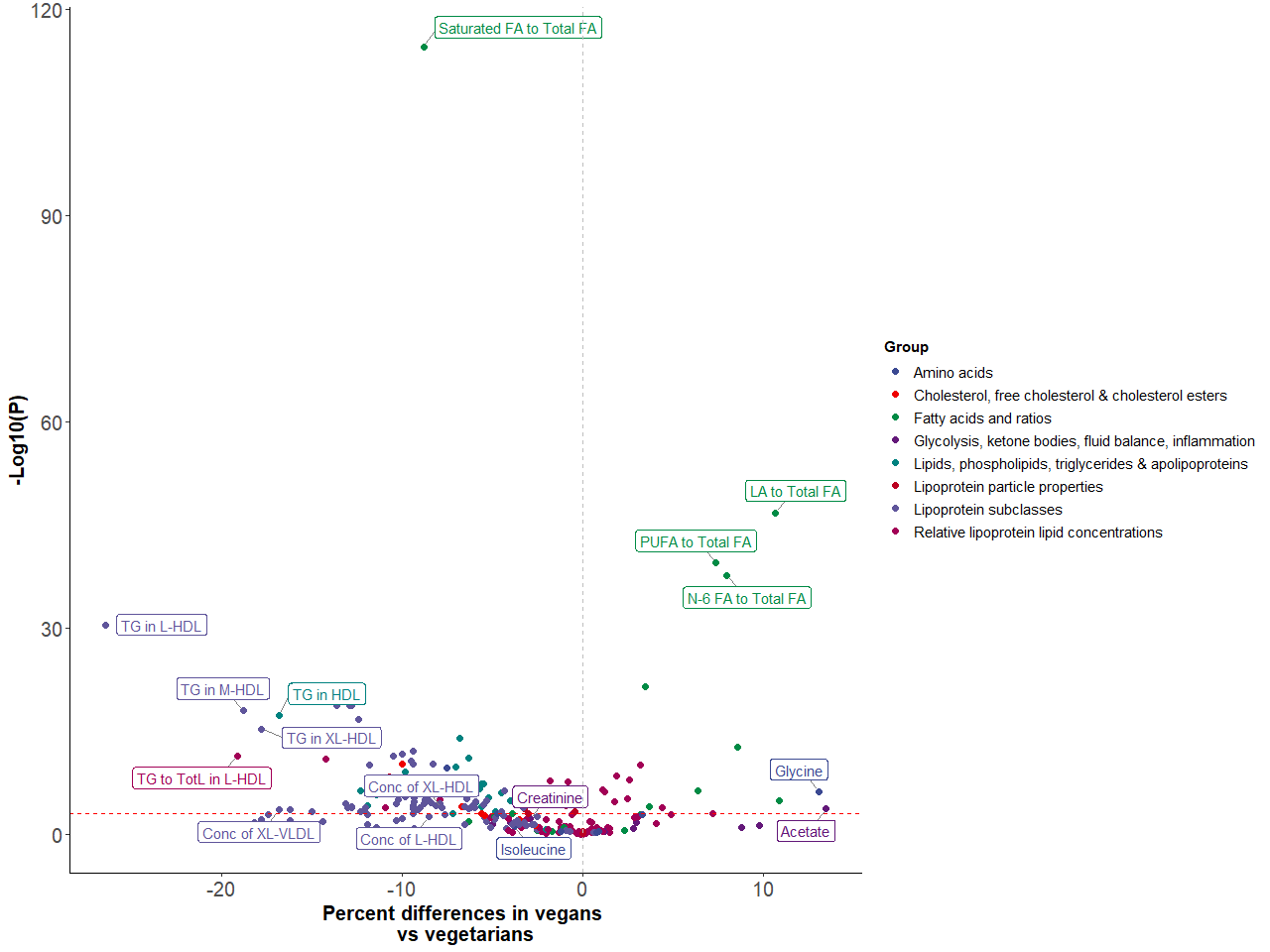

The red dashed line indicates the statistical significance threshold after Bonferroni correction for multiple testing based on the effective number of independent tests (*p* < 0.0010). The grey dashed vertical line marks zero percent difference between diet groups. Chylo: chylomicrons; Conc: concentration; DHA: docosahexaenoic acid; HDL: high-density lipoproteins; L: large; LA: linoleic acid; MUFA: monounsaturated fatty acids; N-3 FA: omega-3 fatty acids; N-6 FA: omega-6 fatty acids; PL: phospholipids; TG: triglycerides; TotL: total lipids; VLDL: very low-density lipoproteins; XL: very large; XXL: Extremely large.

**Supplementary Table 1.** Significant metabolites and their corresponding percent differences from pairwise diet group comparisons in white British and British Indian participants.

Please refer to excel file “Supplementary Table 1.xlsx”, which shows full results from the pairwise comparisons of significant differences in metabolites across six diet groups in white British (using regular meat eaters as the reference group; and vegetarians as the reference group when comparing vegans to vegetarians) and two diet groups in British Indian participants (using meat eaters as the reference group).

The first worksheet summarises all abbreviations used throughout the file. For white British participants, six worksheets present comparisons of low meat eaters, poultry eaters, fish eaters, vegetarians, and vegans versus regular meat eaters, as well as vegans versus vegetarians. For British Indian participants, one worksheet shows the results of comparison between vegetarians and meat eaters.

In all results worksheets, column A lists metabolite group names, with “lower” and “higher” labels underneath to indicate the direction of difference relative to the reference group; column B shows the number of significant metabolites in each category; and column C lists individual metabolite names, along with their corresponding percent differences (in brackets), relative to the reference group.

All percent differences were calculated based on estimates from multivariable-adjusted linear regression models, adjusting for age at recruitment, sex, BMI, alcohol consumption, smoking status, physical activity, geographical region, fasting status, and spectrometer number.

**Supplementary Table 2**. Differences in metabolites by diet group in white British and British Indian populations.

Please refer to excel file “Supplementary Table 2.xlsx”, which shows results for differences in 249 metabolites across six diet groups in white British (worksheet ‘white British participants’) and two diet groups in British Indians (worksheet ‘British Indian participants’).

In both worksheets, column A shows the metabolite name and column B shows the number of participants included in the analyses for the metabolite.

In ‘white British participants’, columns C-H show the estimated geometric mean and its 95% CI for regular meat eaters, low meat eaters, poultry eaters, fish eaters, vegetarians, and vegans, respectively; columns I to M present the percent difference of each metabolite between each diet groups compared with regular meat eaters, based on the estimated geometric means; column N shows the percent difference comparing vegans to vegetarians; columns O shows the *p*-heterogeneity across all diet groups; columns P to T show *p*-value for pairwise comparisons of each diet group against regular meat eaters, column U presents the *p*-value for pairwise comparison of vegetarians and vegans.

In ‘British Indian participants’, columns C and D show the estimated geometric mean and its 95% CI for meat eaters and vegetarians; column E shows the percent difference of each metabolite between vegetarians and meat eaters; and column F shows the corresponding *p*-value.

All estimates were based on multivariable-adjusted linear regression model adjusting for age at recruitment, sex, BMI, alcohol consumption, smoking status, physical activity, geographical region, fasting status, and spectrometer number.

**Supplementary Table 3**. Differences in metabolites by diet group in white British and British Indian populations, excluding BMI from the model.

Please refer to excel file “Supplementary Table 3.xlsx”, which shows results for differences in 249 metabolites across six diet groups in white British (worksheet ‘white British participants’) and two diet groups in British Indians (worksheet ‘British Indian participants’).

The results are presented in the same order as in Supplementary Table 2:

In both worksheets, column A shows the metabolite name and column B shows the number of participants included in the analyses for the metabolite.

In ‘white British participants’, columns C-H show the estimated geometric mean and its 95% CI for regular meat eaters, low meat eaters, poultry eaters, fish eaters, vegetarians, and vegans, respectively; columns I to M present the percent difference of each metabolite between each diet groups compared with regular meat eaters, based on the estimated geometric means; column N shows the percent difference comparing vegans to vegetarians; columns O shows the *p*-heterogeneity across all diet groups; columns P to T show *p*-value for pairwise comparisons of each diet group against regular meat eaters, column U presents the *p*-value for pairwise comparison of vegetarians and vegans.

In ‘British Indian participants’, columns C and D show the estimated geometric mean and its 95% CI for meat eaters and vegetarians; column E shows the percent difference of each metabolite between vegetarians and meat eaters; and column F shows the corresponding *p*-value.

All estimates were based on multivariable-adjusted linear regression model adjusting for age at recruitment, sex, alcohol consumption, smoking status, physical activity, geographical region, fasting status, and spectrometer number.
